# Metabolomic profiling in small vessel disease identifies multiple associations with disease severity

**DOI:** 10.1101/2021.05.13.21257190

**Authors:** Eric L. Harshfield, Caroline J. Sands, Anil M. Tuladhar, Frank-Erik de Leeuw, Matthew R. Lewis, Hugh S. Markus

## Abstract

Cerebral small vessel disease (SVD) is a major cause of vascular cognitive impairment and dementia. There are few treatments, largely reflecting limited understanding of the underlying pathophysiology. Metabolomics can be used to identify novel risk factors in order to better understand pathogenesis and to predict disease progression and severity.

We analysed data from 624 patients with symptomatic cerebral SVD from two prospective cohort studies. Serum samples were collected at baseline and patients underwent MRI scans and cognitive testing at regular intervals with up to 14 years of follow-up. Using ultra-performance liquid chromatography mass spectrometry and nuclear magnetic resonance spectroscopy, we obtained metabolic and lipidomic profiles from 369 annotated metabolites and 54,764 unannotated features and examined their association with respect to disease severity, assessed using MRI SVD markers, cognition, and future risk of all-cause dementia.

Over 100 annotated metabolites were significantly associated with SVD imaging markers, cognition, and progression to dementia. Decreased levels of multiple glycerophospholipids, sphingolipids, and sterol lipids were associated with increased SVD load as evidenced by higher white matter hyperintensities (WMH) volume, lower mean diffusivity normalised peak height (MDNPH), greater brain atrophy, and impaired cognition. Higher levels of several amino acids and nucleotides were associated with higher WMH volume, greater atrophy, and lower MDNPH. Lower baseline levels of carnitines and creatinine were associated with higher annualised change in peak width of skeletonised mean diffusivity (PSMD), and several metabolites, including lower levels of valine, caffeine, and VLDL analytes, were associated with future dementia incidence. Additionally, we identified 1,362 unannotated features associated with lower MDNPH and 2,474 unannotated features associated with increased WMH volume.

Our results show multiple distinct metabolic signatures that are associated with imaging markers of SVD, cognition, and conversion to dementia. Further research should assess causality and the use of metabolomic screening to improve the ability to predict future disease severity and dementia risk in SVD. The metabolomic profiles may also provide novel insights into disease pathogenesis and help identify novel treatment approaches.

## Introduction

Cerebral small vessel disease (SVD) accounts for a quarter of all ischaemic strokes and is the most common pathology underlying vascular cognitive impairment and dementia. ^1^ SVD is characterised by typical radiological features seen on brain MRI including lacunes, white matter hyperintensities (WMH), cerebral microbleeds, diffuse ultrastructural changes that can be detected using diffusion tensor imaging (DTI), and brain atrophy. Despite its importance, there are few effective treatments for delaying disease progression. A major reason for this is limited understanding of the disease pathogenesis. Furthermore, although it is a major cause of dementia, only a proportion of patients with radiological SVD progress to cognitive impairment.^2^ Once effective treatments become available, predicting which patients are at elevated risk will become clinically important, and better markers of disease progression are therefore required.^3^

Metabolomics, the high-throughput identification and quantification of small molecules in biological samples, offers the potential to both identify novel disease mechanisms and develop better predictive markers.^4^ Metabolomics assays surpass standard chemistry techniques for the purposes of comprehensive metabolome measurement^5^ since they are capable of precise analysis of hundreds or even thousands of metabolites. ^6^ This allows detailed characterisation of metabolic phenotypes, enabling characterisation of metabolic arrangements underlying disease pathogenesis, discovery of new therapeutic markers, and identification of novel biomarkers to diagnose and monitor disease. ^6^ Metabolomics has been applied successfully in a number of cardiovascular and neurological diseases,^7,8^ but there have been few studies in SVD.

Ultra-performance liquid chromatography mass spectrometry (UPLC-MS) and nuclear magnetic resonance (NMR) spectroscopy are effective analytical techniques for detecting and measuring chemical constituents within blood samples. In this analysis we obtained baseline metabolomics profiles from 624 patients with symptomatic MRI-confirmed SVD and up to 14 years of follow-up. We examined associations between metabolites and disease severity, assessed using both MRI disease markers and cognitive parameters. We also evaluated relationships between metabolites and future risk of all-cause dementia.

## Materials and methods

### Data sources

We analysed individual participant data from two studies involving patients with symptomatic SVD: first, St George’s Cognition and Neuroimaging in Stroke (SCANS), a longitudinal study of cognitive impairment in 121 patients with moderate to severe symptomatic SVD; ^2,9^ and second, the Radboud University Nijmegen Diffusion Tensor and Magnetic Resonance Imaging Cohort (RUN-DMC), a prospective cohort study from the Netherlands of 503 individuals aged between 50-85 years with symptomatic SVD.^10^ SCANS participants had multimodal MRI and cognitive tests performed at baseline and at years 1, 2, and 3, as well as 5-year follow-up for dementia, and RUN-DMC participants had MRI, cognitive, and clinical assessments performed at baseline and at years 5, 9, and 14, with 14 years of follow-up for dementia. Both studies recorded information from each participant on a range of demographics and vascular risk factors, including sex, age, ethnicity, body mass index, smoking status, diabetes status, systolic and diastolic blood pressure, hypertension status, and hypercholesterolemia status. Follow-up data on dementia incidence was available for all 121 patients from SCANS and 501 patients from RUN-DMC.

### Metabolomics data

Serum samples collected at baseline from 624 participants from the SCANS and RUN-DMC cohorts were analysed using UPLC-MS and proton ^1^H NMR spectroscopy. Full analytical details, following previously described sample preparation, analytical, and quality control (QC) procedures,^11–13^ are provided in the **Supplementary Methods**. For each assay, samples were analysed in a randomised order demonstrating no correlation or other relationship with study design variables, precluding any confounding effect of analysis order. To facilitate quality assessment and pre-processing, a pooled QC sample was prepared by combining equal parts of each study sample and analysed periodically among study sample analyses. For UPLC-MS only, a series of QC sample dilutions was created (10 x 100%, 5 x 80%, 3 x 60%, 3 x 40%, 5 x 20%, 10 x 1%) and analysed at the start and end of each set of sample analyses.

NMR and UPLC-MS assays were applied to maximise coverage of a broad range of metabolite classes including lipophilic, hydrophilic, small and macromolecular analytes and processed to include both global profiling and targeted extraction datasets (**Table 1**). Global profiling provides a comprehensive analysis of all measurable metabolites in a sample but results in datasets with large numbers of variables per analyte, the identities of which are typically unknown. In contrast, by targeted extraction of a pre-defined set of metabolites, pre-annotated datasets are immediately more interpretable but are limited in coverage to those metabolites in the pre-defined set.

**Table 1.** Metabolites analysed using each metabolic profiling assay.

| Technology platform | Metabolic profiling assay | No. features (global profiling datasets) | No. annotated metabolites (targeted extraction datasets) | Metabolome/lipidome coverage | No. participants |
| --- | --- | --- | --- | --- | --- |
| UPLC-MS | HILIC+ | 5,729 | 29 | Hydrophilic metabolites including carnitine, betaine, warfarin, caffeine, cotinine, metform, trimethylamine-N-oxide (TMAO), proline, creatine, cytosine | SCANS: 83<br>RUN-DMC: 376<br>Total: 459 |
|  | Lipid RPC- | 4,336 | 31 | Lipophilic metabolites including bilirubin, fatty acids, lysophosphatic acids, lysophosphocholines, lysophosphoethanolamines | SCANS: 101<br>RUN-DMC: 447<br>Total: 548 |
|  | Lipid RPC+ | 7,407 | 190 | Lipophilic metabolites including carnitines, cholesteryl esters, ceramides, cholesterol, diglycerides, lysophosphocholines, lysophosphoethanolamines, monoacylglycerols, phosphocholines, phosphethanolamines, sphingomyelins, triglycerides | SCANS: 101<br>RUN-DMC: 456<br>Total: 557 |
| NMR | Standard 1D | 18,646 | 14 (IVDr BI-QUANT) | Small molecule metabolites including creatinine, trimethylamine-N-oxide (TMAO), alanine, creatine, glutamine, histidine, isoleucine, tyrosine, valine, lactic acid, acetoacetic acid, glucose | SCANS: 115<br>RUN-DMC: 494<br>Total: 609 |
|  |  |  | 105 (IVDr BI-LISA) | Lipoprotein subfractions including subtypes of cholesterol, phospholipids, triglycerides, and apolipoproteins | SCANS: 105<br>RUN-DMC: 430<br>Total: 535 |
|  | CPMG | 18,646 | N/A | Small molecule metabolites | SCANS: 111<br>RUN-DMC: 451<br>Total: 562 |
| **Overall** |  | **54,764** | **369** |  | **SCANS: 121**<br>**RUN-DMC: 503**<br>**Total: 624** |

UPLC-MS was applied with two chromatographic techniques: hydrophilic interaction chromatography (HILIC), for the separation of hydrophilic analytes (i.e. polar and charged metabolites) and reverse-phase chromatography (RPC) for the separation of lipophilic analytes (i.e. complex and neutral lipids). When coupled to positive and/or negative mode ionisation the following datasets were produced: lipid positive (lipid RPC+), lipid negative (lipid RPC-) and HILIC positive (HILIC+). NMR assays comprised a standard one-dimensional (1D) NMR profile experiment with water pre-saturation using the 1D-NOESY pre-sat pulse sequence for characterisation of small and macro-molecular metabolites and an additional spin-echo experiment using the 1D Carr-Purcell-Meiboom-Gill (CPMG) pre-sat pulse sequence for saturation of macromolecules signals.

For generation of global profiling UPLC-MS datasets, untargeted peak detection was performed using Progenesis QI (Waters Corp., Manchester, UK). For targeted extraction, an in-house algorithm developed at the National Phenome Centre (peakPantheR, github.com/phenomecentre/peakPantheR) was used to fit pre-defined UPLC-MS signals with semi-automated (manually validated) extraction of known chemical species across the three assays. For NMR, targeted extraction was performed using the in vitro diagnostics platform (IVDr) from Bruker Biospin (www.bruker.com) generating quantified measurements of both lipoprotein subclasses (BI-LISA) and small molecules (BI-QUANT).

For all datasets, pre-processing and QC was performed using the nPYc-Toolbox^14^ according to previously published criteria. ^11,13^ Metabolite intensity values on the lipid RPC+ and lipid RPC-platforms were corrected for run-order and batch-related intensity drifts by applying LOESS regression fitted to the pooled QC samples. Run-order and batch correction were not necessary for the metabolites measured on the HILIC+ platform. Only features/metabolites measured with high analytical quality (RSD in pooled QC<30%, dilution series Pearson correlation to dilution factor>0.7, RSD in study samples>1.1* RSD in pooled QC) were retained. For the global profiling datasets this resulted in 5,729 features for HILIC+, 4,336 features for lipid RPC-, and 7,407 features for lipid RPC+. For the targeted extraction UPLC-MS datasets, a total of 250 unique and known chemical species passed QC across the three assays (29 on HILIC+, 31 on lipid RPC-, and 190 on lipid RPC+). For NMR global profiling data, after removal of uninformative spectral regions, 18,646 features were available in both standard 1D and CPMG NMR datasets. Quantification using the Bruker BI-QUANT algorithm resulted in automated quantification of 27 small molecules, of which 14 passed the feature selection criteria; the remaining 13 metabolites were not detected or were not present in sufficient concentrations to be measured accurately. Application of the BI-LISA algorithm resulted in automated quantification of 105 lipoprotein subclasses. Across all assays, discrepancies in final sample numbers available for analysis (**Table 1**) result from insufficient sample volume for data acquisition, sample compromised during acquisition or sample exclusion owing to data not meeting stringent quality control criteria (**Supplementary Materials**). To ensure approximately normal distributions, a generalised log transformation was applied to all features/metabolites and the values were rescaled using mean-centring and dividing by the standard deviation of each metabolite across participants.

### MRI and clinical endpoints

Our primary MRI endpoint was baseline mean diffusivity normalised histogram peak height measured within normal appearing white matter voxels (MDNPH), a diffusion tensor imaging (DTI) marker that has previously been shown to be correlated with, and predictive of, the degree of cognitive impairment. ^2,15^ A reduction in MDNPH corresponds with increasing mean diffusivity. Secondary MRI endpoints that we examined were baseline cerebral microbleed count, lacune count, WMH (expressed as the percentage of WMH volume out of the total brain volume), total brain volume, and peak width of skeletonised mean diffusivity (PSMD), an alternative DTI marker that has been shown to be robust and highly sensitive. ^16^ Our primary clinical endpoint was conversion to dementia, which was diagnosed using the 5^th^ edition of the Diagnostic and Statistical Manual of Mental Disorders (DSM-5) definition for major neurocognitive disorder. Secondary clinical endpoints that we examined consisted of: cognition, assessed by a global cognition score as well as scores for the executive function and processing speed domains; and disability, assessed by the Barthel index, which is used to measure performance on Activities of Daily Living (ADL). We orientated each outcome so that higher values corresponded to increased cognitive decline (e.g. instead of total brain volume we analysed brain atrophy as its inverse). We analysed cerebral microbleed and lacune counts both as continuous and binary variables (i.e. presence or absence of microbleeds or lacunes). A further endpoint that we included was a simple MRI score that accounted for presence of microbleeds, number of lacunes, and WMH volume (Fazekas score), which has been shown to improve prediction of dementia in SVD patients. ^9^ To obtain comparable effect sizes across outcomes, values for each outcome were rescaled using mean-centring and dividing by the standard deviation across participants. A description of these endpoints is provided in **Supplementary Table 1**.

### Statistical analyses

We performed cross-sectional analyses examining the association of baseline MRI markers, cognition, and disability data per 1-SD higher metabolite levels measured at baseline. We constructed linear regression models for continuous outcomes and logistic regression models for binary outcomes, with adjustment for cohort, baseline age, and sex. To evaluate the relationship of metabolites with changes in MRI parameters and cognition over time, we calculated an annualised change in values for each outcome, based on the difference in values between the baseline and latest time point divided by the amount of follow-up time that had elapsed. We then ran linear regression models examining the association of annualised change in MRI markers, cognition, and disability per 1-SD higher metabolite levels. We also performed longitudinal analyses to determine whether metabolites measured at baseline predict long-term conversion to dementia, for which we constructed Cox proportional-hazards regression models adjusted for cohort, age, and sex to assess the association of conversion to dementia per 1-SD higher metabolite levels.

Analyses were conducted using R version 4.0.2 (R Core Team, 2020). To account for multiple testing comparisons, we used a false discovery rate (FDR) threshold of *q* < 0.05 to identify significant associations for each outcome measure. Two-sided *P*-values and 95% confidence intervals are presented.

### Sensitivity analyses

As a sensitivity analysis, we conducted analyses at baseline separately within each cohort for significantly associated metabolites to compare the magnitude and direction of associations across cohorts. We also examined significantly associated metabolites to determine whether the associations of metabolites with imaging markers and cognition were modified by relevant risk factors. We repeated the cross-sectional analyses with further adjustment for diabetes status, hypertension status, and hypercholesterolemia status.

### Data availability

The raw metabolomics data described in this study were generated at the Medical Research Council National Institute for Health Research (MRC-NIHR) National Phenome Centre. Derived data supporting the findings of this study are available from the corresponding author on request.

## Results

### Patient characteristics

In this study we analysed individual participant data from 624 patients with symptomatic SVD. The majority of participants were male (58%) and Caucasian (91%), with a mean (SD) age of 66.5 (9.2) years (**Supplementary Table 2**). Compared to participants from RUN-DMC, SCANS participants were on average 4.4 years older, came from more diverse ethnic backgrounds (23% Caribbean and 6% African in SCANS), had higher rates of hypertension and hypercholesteremia, and had more severe SVD as indicated by increased WMH volume.

### Associations with baseline imaging parameters

We obtained measurements for 369 annotated metabolites and lipoprotein subclasses measured on five different metabolomics platforms that employed both UPLC-MS and NMR (**Table 1**). We analysed the association of these metabolites with a range of MRI markers, indicators of cognition and disability, and conversion to dementia (**Supplementary Table 1**).

In cross-sectional analyses adjusted for cohort, baseline age, and sex, lower serum levels of 34 sphingolipids (including sphingomyelins and ceramides) were associated with lower MDNPH, higher WMH volume, greater brain atrophy, and impaired cognition (**Figure 1a, Supplementary Table 3**). Lower levels of 30 glycerophospholipids (including phosphatidylcholines and lysophosphatidylcholines) were also associated with lower MDNPH, greater brain atrophy, and impaired cognition. Higher levels of 7 amino acids and nucleotides (N^1^-Acetylspermidine, N-Acetylputrescine, isoleucine, creatinine, creatine, cytosine, and 5’-Methylthioadenosine) were associated with lower MDNPH, higher WMH volume, and greater brain atrophy (**Figure 1b, Supplementary Table 3**). Lower levels of bilirubin were associated with impaired cognition. Higher levels of caffeine were associated with greater brain atrophy but also with improved cognition.

**Figure 1.**
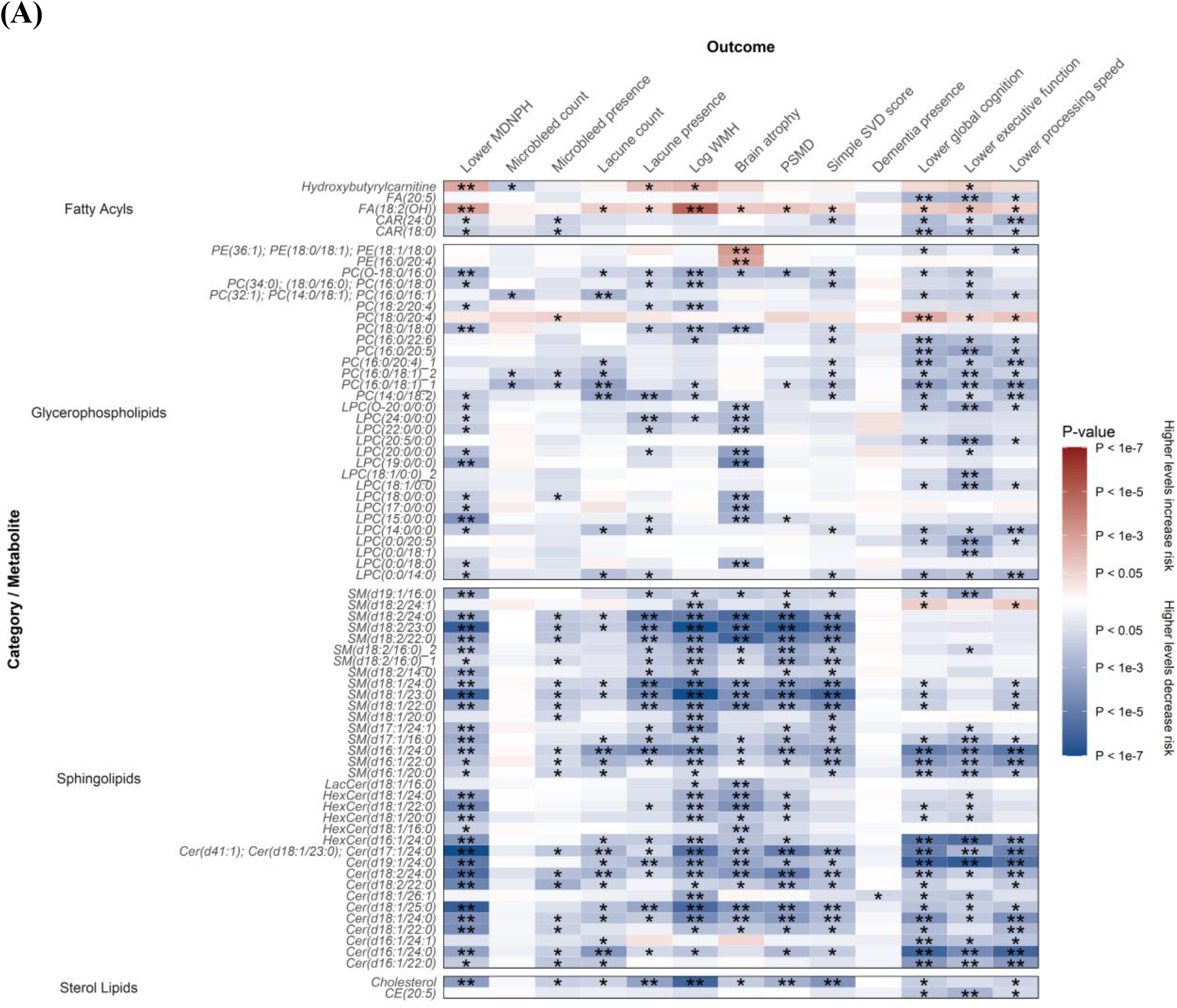
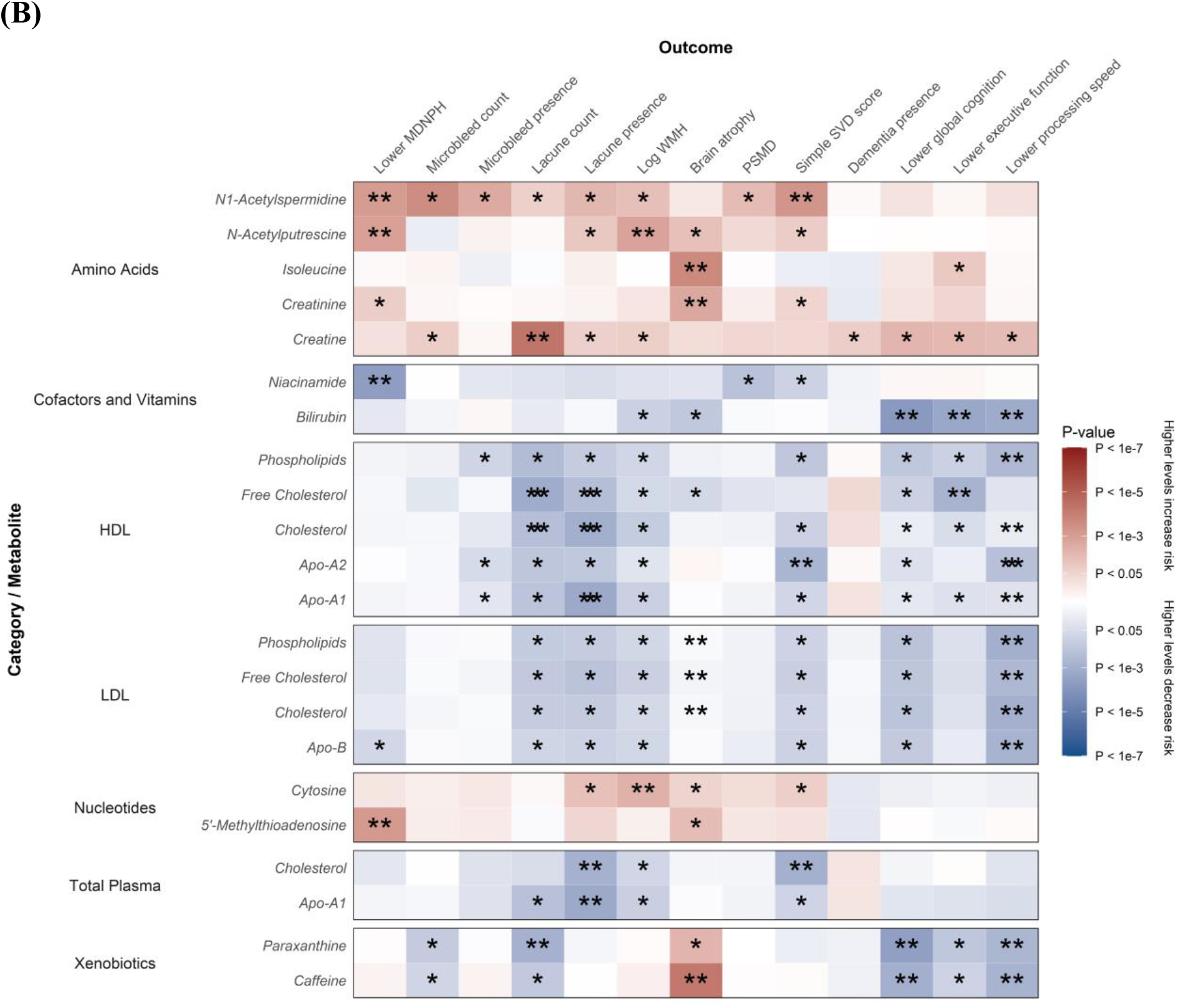
Association of MRI markers and cognition parameters at baseline per 1-SD higher metabolite levels. (**A**) Lipids. (**B**) Lipoproteins and small molecules. Beta estimates and *P*-values were obtained from linear or logistic regression models adjusted for baseline age, sex, and cohort. Colours show magnitude and direction of *P*-value for association of metabolite with each outcome (red indicates positive association and blue indicates inverse association). Asterisks indicate significance: * *P* < 0.05; **FDR *q* < 0.05.

### Longitudinal analyses of progression of MRI parameters and cognition and of incident dementia

We also analysed the association of metabolites at baseline with annualised change in levels of imaging markers and cognition (**Figure 2, Supplementary Table 4**). Lower levels of four carnitines and creatinine were associated with higher annualised change in PSMD, and lower levels of 23 lipoprotein analytes in IDL, LDL, VLDL, and total plasma were associated with higher annualised change in impaired executive function. Higher levels of creatine and glucose were associated with increased annualised change in number of lacunes.

**Figure 2.**
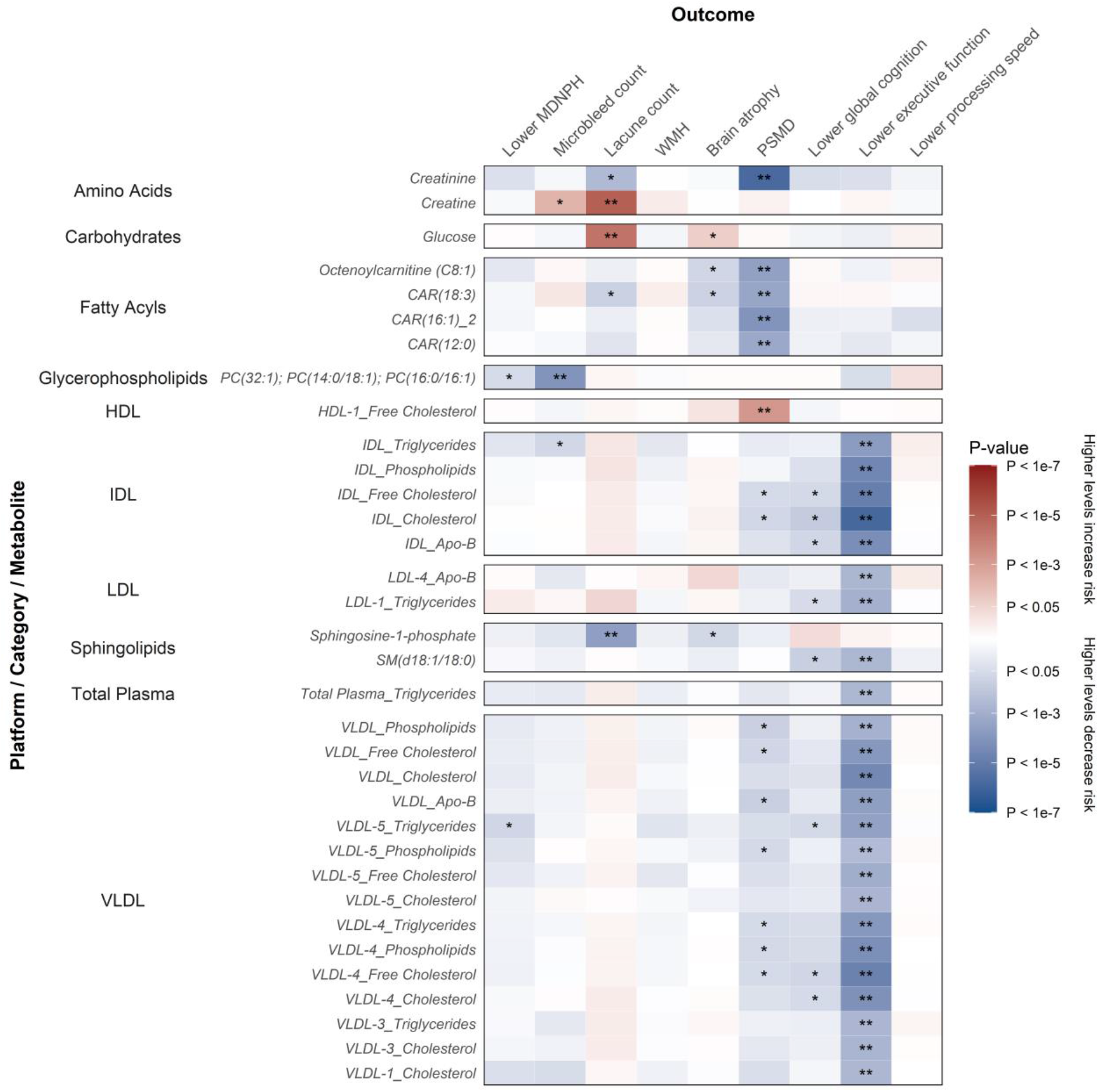
Association of annualised change in MRI markers and cognition parameters per 1-SD higher metabolite levels. Annualised change in outcomes were calculated as difference in values between baseline and latest time point divided by amount of follow-up time. Beta estimates and *P*-values were obtained from linear or logistic regression models adjusted for baseline age, sex, and cohort. Colours show magnitude and direction of *P*-value for association of metabolite with each outcome (red indicates positive association and blue indicates inverse association). Asterisks indicate significance: * *P* < 0.05; **FDR *q* < 0.05.

When accounting for long-term follow-up in time-to-event analyses, future incidence of dementia was associated with several metabolites, including lower levels of valine, caffeine, and VLDL analytes, and higher levels of urocanate, lipoprotein analytes in HDL and LDL, and creatine (**Figure 3, Supplementary Table 5**). These associations were suggestive (*P* < 0.05) but not statistically significant (FDR *q* < 0.05) after correcting for multiple testing.

**Figure 3.**
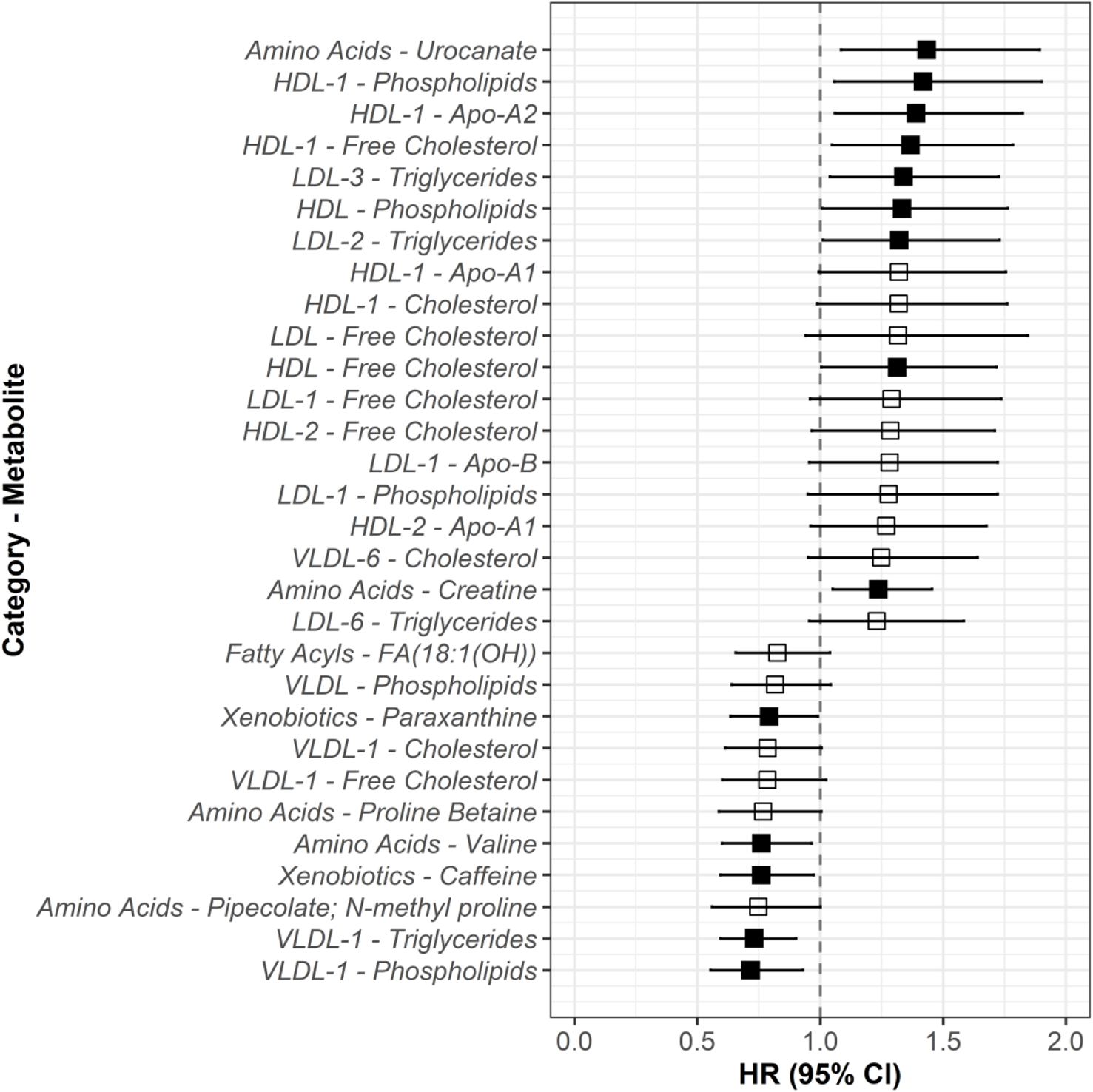
Adjusted hazard ratios for dementia per 1-SD higher metabolite levels. Analyses were adjusted for baseline age, sex, and cohort. Hollow squares indicate associations that were not statistically significant ; filled squares indicate associations significant at *P* < 0.05.

### Sensitivity analyses

Analyses conducted separately within each cohort showed that the directions of association were mostly consistent between SCANS and RUN-DMC, but there were some differences in the magnitudes of the associations (**Supplementary Fig. 1, Supplementary Table 6**). Lower levels of multiple glycerolipids (triglycerides and diglycerides) were associated with lower MDNPH and impaired cognition in SCANS participants, with no evidence of an association in RUN-DMC participants. Lower levels of multiple glycerophospholipids and sphingolipids were associated with lower MDNPH and impaired cognition in both SCANS and RUN-DMC, but the specific lipids that reached statistical significance within each lipid class varied. Lower levels of many of these sphingolipids were also associated with increased WMH volume, greater atrophy, and higher PSMD in RUN-DMC participants, but not in SCANS participants.

In analyses adjusted for additional vascular risk factors (diabetes, hypertension, and hypercholesterolemia status), many of the associations attenuated and were no longer statistically significant (**Figure 4, Supplementary Table 7**). Only one amino acid (creatine), one fatty acyl [FA(18:2(OH))], five glycerophospholipids, 17 sphingolipids, one sterol lipid (cholesterol), one lipoprotein analyte (Apo-A2 in HDL-4), and two xenobiotics (paraxanthine and caffeine) remained statistically significant after this further adjustment.

**Figure 4.**
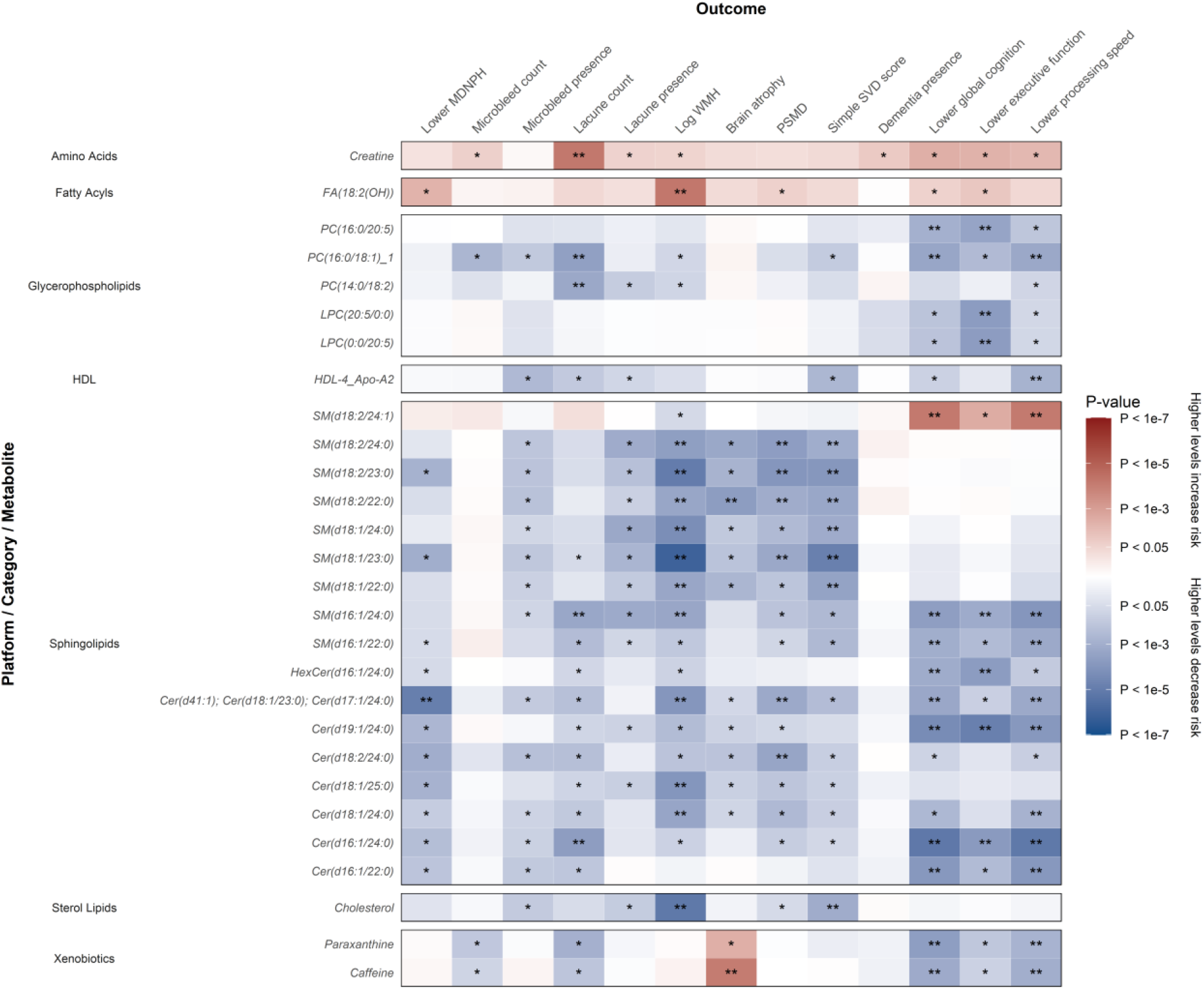
Association of MRI markers and cognition parameters at baseline per 1-SD higher metabolite levels with further adjustment for relevant risk factors. Beta estimates and *P*-values were obtained from linear or logistic regression models adjusted for cohort, baseline age, sex, diabetes status, hypertension status, and hypercholesterolemia status. Colours show magnitude and direction of *P*-value for association of metabolite with each outcome (red indicates positive association and blue indicates inverse association). Analyses were restricted to metabolite-outcome associations that were significant in the primary analysis (**Figure 1**). Asterisks indicate significance: * *P* < 0.05; **FDR *q* < 0.05.

### Analyses of global profiling datasets

To provide a more global overview, in addition to analyses conducted on the targeted extraction datasets, analyses were also conducted on the unannotated, global profiling datasets. These analyses also revealed statistically significant associations with a number of features. From a total of 54,764 measured features, after correcting for multiple testing using an FDR threshold of *q* < 0.05, we identified 1,362 features associated with lower MDNPH, 2,474 features associated with increased WMH volume, and 1,533 features associated with executive function (**Supplementary Tables 8** and **9**). Despite the larger number of features measured using NMR, a greater proportion of the significant associations were with features derived from UPLC-MS datasets, resulting from increased depth of coverage (UPLC-MS assays weighted to lipids, NMR weighted to small molecules) and the higher degree of redundancy in the NMR global profiling data (multiple features derived from the same underlying metabolite). Similar to the analyses conducted in the smaller set of annotated metabolites, there were no significant associations of features with microbleed count or conversion to dementia in the global profiling datasets.

## Discussion

In this comprehensive metabolomics profiling study of over 600 individuals with MRI-confirmed SVD, we identified over 100 annotated metabolites that are significantly associated with SVD imaging markers, cognition, and progression to dementia. We found that decreased levels of multiple glycerophospholipids, sphingolipids, and sterol lipids are associated with increased SVD load as evidenced by higher WMH volume, lower MDNPH, and greater atrophy, as well as with impaired cognition. We also found that higher levels of several amino acids and nucleotides are associated with higher WMH volume, greater atrophy, and lower MDNPH. Several metabolites, including lower levels of valine, caffeine, and VLDL analytes, are associated with future dementia incidence.

The associations with glycerophospholipids, sphingolipids, and sterol lipids were particularly notable. Previous metabolomics studies have shown associations of lower levels of ceramide ratios with fewer number of cerebral microbleeds^17^ and increased risk of incident dementia.^18^ In the present study, lower levels of serum sphingomyelins and ceramides were associated with higher WMH volume, greater brain atrophy, and impaired cognition in baseline analyses adjusted for cohort, age, and sex. The associations for 16 sphingolipids remained significant even after further adjustment for diabetes, hypertension, and hypercholesterolemia status, suggesting that these associations are not explained by other risk factors and may have causal mechanisms. Only one sphingomyelin had a statistically significant association with executive function when assessing the annualised change in metabolite levels, suggesting that the absolute levels of the metabolites at baseline are more relevant in evaluating their effects on SVD and cognition than how those levels change over time.

Demyelinating diseases such as multiple sclerosis cause neuroinflammation, which can result in damage to the myelin sheath. Inflammation has also been proposed to play a role in the progression of SVD, ^19^ and metabolites could be implicated in the causal pathway. Previous studies have shown that patients with multiple sclerosis and other demyelinating diseases have increased levels of sphingomyelins and ceramides in cerebrospinal fluid. ^20,21^ However, these ceramides and sphingomyelins have also been implicated in non-neurological conditions such as heart failure.^22^

Linoleic acid [FA(18:2(OH))] is an essential omega-6 fatty acid obtained from plant sources. Diets rich in linoleic acid and other omega-6 fatty acids inhibit the metabolic formation of omega-3 polyunsaturated fatty acids, which can lead to a deficit of eicosapentaenoic acid (EPA)^23^ and is associated with reduced brain volume, impaired cognition, and accelerated progression to dementia.^24^ Our study showed that increased levels of linoleic acid were associated with higher WMH volume.

Another finding from our study was that increased caffeine consumption (i.e. higher levels of caffeine in serum) was associated with lower total brain volume but improved cognition, particularly processing speed, and decreased risk of dementia. Numerous systematic reviews have demonstrated the positive benefits of caffeine consumption, ^25^ but studies have also shown that coffee consumption is associated with increased risk of Alzheimer’s disease,^26,27^ though there is no evidence of a causal relationship of coffee consumption with small vessel disease or other ischaemic stroke subtypes.^28^ One explanation for our findings is that caffeine can be associated with short-term improvement in cognitive functioning but that long-term consumption is associated with chronic brain atrophy.

A previous analysis of eight prospective cohort studies found that increased levels of isoleucine, creatinine, and VLDL lipoprotein subclasses were associated with lower risk of dementia.^29^ In our study, increased levels of isoleucine and creatine were associated with increased brain atrophy, and VLDL analytes were associated with lower risk of incident dementia.

Our findings have several important clinical implications. First, they may provide novel insights into pathogenic mechanisms underlying SVD; it is possible that modifying levels of specific metabolites could help reduce the risk of cognitive decline and dementia in patients with SVD. Dietary interventions or novel therapies could improve long-term outcomes for SVD patients. Second, a metabolomics panel based on these associations could be developed for clinicians to predict which patients are most likely to progress to more severe forms of dementia and offer personalised treatment. Third, exploration of the broader metabolic profiles derived from our investigation show promise for the discovery and identification of additional markers yielding greater mechanistic insight to the relevant phenotypes.

The strengths of our study include the fact that the metabolites, indicators of cognitive function, and brain MRI markers were measured together at baseline, with MRI and cognitive data also available at multiple timepoints, and with long-term prospective follow-up of 5-14 years. Second, the metabolites were measured using a robust, highly accurate, validated analytical approach with quality control measures. Third, we conducted sensitivity analyses separately within each cohort and with adjustment for additional relevant risk factors to assess potential mediators.

The differences in the magnitudes of the associations between cohorts is likely because RUN-DMC had a larger population and a wider range of disease, whereas SCANS was a much smaller population and was more homogeneous, with all patients having moderate or severe SVD on MRI. The reason for an association of triglycerides and diglycerides with lower MDNPH and impaired cognition in SCANS but not in RUN-DMC could be because participants in RUN-DMC had fasted overnight before blood samples were taken, which can have a significant impact on triglyceride levels. ^30^

Our study also has limitations. First, despite the fact that this is one of the largest metabolomics studies on SVD to date, the sample sizes of the studies were still modest, which reduced the power to detect associations. Second, the large number of statistical tests conducted meant that some associations may have been biologically and clinically meaningful but did not reach the threshold for statistical significance after correction for multiple testing. However, we applied an FDR correction to reduce the likelihood of identifying false positives. Third, we were unable to determine whether the associations that we identified were causal, as we did not have access to genetic instruments for the measured metabolites that could be used in a Mendelian randomization analysis. The metabolites could be on the causal pathway but secondary to tissue damage caused by demyelination. ^31^ Patients in SCANS were not assessed for multiple sclerosis and myelin loss was not measured so we were unable to evaluate this, and further mechanistic and longitudinal studies are needed. However, even if changes in metabolite levels do not directly cause cognitive decline or dementia, they could still be useful predictors of these conditions. Fourth, the metabolites were measured in blood serum rather than cerebrospinal fluid, which is considered better suited for measurement of sensitive biomarkers of neurological and cognitive decline, ^32^ although studies have shown similar changes in affected pathways for metabolites measured in blood and CSF. ^33^ However, serum biomarkers are clinically useful as serum is much less invasive to collect from patients. Fifth, we did not examine ratios of metabolites, which can reveal additional insights into metabolic pathways^34^ and should be examined in follow-up analyses. Finally, the study was conducted in patient populations with symptomatic SVD and may not be generalisable to other contexts.

In conclusion, we provide consistent evidence that multiple serum metabolites are associated with SVD severity on MRI, cognitive decline, and incident dementia in patients with cerebral SVD. Further research should be conducted to identify if these associations are causal and could be used to improve the ability of clinicians to predict the rate of progression and severity of onset of lacunar stroke and vascular dementia, and for researchers to develop novel treatment approaches for patients at increased risk of these conditions.

## Abbreviations

ADL: Activities of Daily Living
BI-LISA: Bruker IVDr Lipoprotein Subclass Analysis
BI-QUANT: Bruker IVDr automated quantification of small molecule metabolites
DTI: diffusion tensor imaging
DSM: Diagnostic and Statistical Manual of Mental Disorders
FDR: false discovery rate
LOESS: locally estimated scatterplot smoothing
MDNPH: mean diffusivity normalised histogram peak height measured within normal appearing white matter voxels
NMR: nuclear magnetic resonance
UPLC-MS: ultra-performance liquid chromatography mass spectrometry
PSMD: peak width of skeletonised mean diffusivity
RUN-DMC: Radboud University Nijmegen Diffusion Tensor and Magnetic Resonance Imaging Cohort
SCANS: St George’s Cognition and Neuroimaging in Stroke
SD: standard deviation
SVD: small vessel disease
WMH: white matter hyperintensities

