## Supplementary Materials for "Metabolomic profiling in small vessel disease identifies multiple associations with disease severity"

### Supplementary Material

**Supplementary Methods**

Metabolomics sample handling

Metabolomics analytical methods and coverage

**Supplementary Tables**

Supplementary Table 1. Outcomes included in this analysis

Supplementary Table 2. Characteristics of included studies

Supplementary Table 3. All results for association of MRI markers and cognition parameters at baseline per 1-SD higher metabolite levels

Supplementary Table 4. All results for association of annualised change in MRI markers and cognition parameters per 1-SD higher metabolite levels

Supplementary Table 5. All results from Cox proportional-hazards regression models for dementia per 1-SD higher metabolite levels

Supplementary Table 6. All results for association of MRI markers and cognition parameters at baseline per 1-SD higher metabolite levels by cohort

Supplementary Table 7. All results for association of MRI markers and cognition parameters at baseline per 1-SD higher metabolite levels with further adjustment for relevant risk factors

Supplementary Table 8. Summary of association of global profiling features with imaging and cognitive parameters

Supplementary Table 9. Significant results from association of global profiling features with imaging and cognitive parameters

**Supplementary Figures**

Supplementary Figure 1. Association of MRI markers and cognition parameters at baseline per 1-SD higher metabolite levels by cohort

### Supplementary Methods

#### Metabolomics sample handling

Upon receipt, all samples were unpacked from their shipping container and stored at -80°C. Samples were sorted into batches of 80 using metal racks in contact with dry ice, ensuring the samples were maintained at a temperature of -40°C or below at all times. The sample sorting order was determined by orthogonalization with respect to the study design provided to the National Phenome Centre (NPC). Samples were returned to -80°C until needed for sample aliquoting.

For aliquoting, samples were thawed at 4°C overnight in metal racks providing an equal rate of sample thawing, and the liquid contents were transferred to a 96-well plate. Each plate was centrifuged to remove particulate matter, and the supernatant was aspirated and dispensed across dedicated 96-well plates for each assay. Plates were heat-sealed and returned to -80°C until needed for assay-specific sample preparation.

#### Metabolomics analytical methods and coverage

All NPC assays include repeat measurements of a set of reference samples in addition to any assay-specific calibration samples outlined in the publications referenced below. Pooled QC samples consist of a pool of all samples for each matrix in the study, and, when acquired periodically throughout the analysis, allow for correction of longitudinal drift in analytical measurement as well as the calculation of quality metrics for each feature detected. For UPLC-MS only, an additional series of QC sample dilutions was created (10x 100%, 5x 80%, 3x 60%, 3x 40%, 5x 20%, 10x 1%) in order to assess linearity of response. Long-Term Reference (LTR) samples are a pool of samples of an identical matrix (serum) external to the study which is used internally for quality control purposes.

##### Nuclear Magnetic Resonance Spectroscopy

Acquisition of NMR profiles was conducted according to previously published protocols.[11] In brief, 300 µL of serum sample were mixed with 300 µL of $H_2O$:$D_2O$ buffer containing 75 mM of $Na_2HPO_4$, 4.6 mM of TSP (3-(trimethylsilyl)-2,2,3,3-tetradeuteropropionic acid) and 6.2 mM of $NaN_3$ at pH 7.4. Samples were prepared into 4 inch length, 5 mm outer diameter NMR tubes in 96-well plate formation using a Gilson 215 liquid handling robot (Gilson S.A.S,

France). Each rack, containing 80 study samples, two pooled QC and two LTR samples, was transferred to a refrigerated SampleJet automated sample manager (Bruker Corporation, Bruker) thermostated at 6°C. $^{1}$H-NMR spectra were produced for each sample by performing a standard one-dimensional (1D) profile experiment, a two-dimensional J-resolved experiment, and a spin-echo experiment using the $^{1}$D-CPMG presat pulse sequence on a Bruker 600 Avance III HD spectrometer with TopSpin 3.6 and ICON NMR. Phasing, baseline correction and calibration were carried out in automation after each experiment using TopSpin 3.6, with serum samples calibrated to the anomeric proton signal of a-glucose at 5.233 ppm. Spectra that passed the analytical checks were aligned to a common reference scale, running from 10 to -1 ppm, and interpolated onto a common 20,000 point grid.

Spectral quality was assessed using a set of in-house scripts following quality criteria as previously described:[11,14]

- Line width of less than 1.4 Hz
- Quality of water-suppression (the residual water signal did not impinge on the surrounding spectrum)
- Even baseline signal (the baseline was even and flat across the entire spectrum)
- Accurate chemical shift referencing

Quality assessment was performed daily, and any spectrum failing any of the above tests was immediately rerun. If the sample did not pass a second time, it was excluded from further analysis.

Lipoprotein and small molecule quantification were generated by the in vitro diagnostics platform (IVDr) from Bruker Biospin (www.bruker.com), using the BI-LISA and BI-QUANT protocols respectively.

#### Ultra-Performance Liquid Chromatography - Mass Spectrometry

Mass Spectrometry (MS) provides a highly sensitive platform for generating quantifications of small molecules, and when combined with the appropriate Ultra-Performance Liquid Chromatography (UPLC) provides a wide coverage of biologically relevant metabolic classes. Sample preparation for acquisition of profiling datasets by UPLC-MS was conducted in batches of 80, daily, to minimise the effect of sample aging.

Serum samples were prepared as previously described for the separation of lipophilic analytes (e.g. complex and neutral lipids) by reversed-phase chromatography (lipid RPC) and the separation of hydrophilic analytes (e.g. polar and charged metabolites) by hydrophilic interaction liquid chromatography (HILIC).[13] Briefly, 50 μL aliquots were taken from each sample and the pooled QC and diluted 1:1 (v/v) with ultrapure water. Protein was removed by addition of organic solvent (diluted sample/isopropanol in 1:4 (v/v) ratio for lipid RPC profiling and diluted sample/acetonitrile in 1:3 (v/v) ratio for HILIC profiling). Mixtures of method-specific authentic chemical standards were added at the dilution step (HILIC) or the protein precipitation step (lipid-RPC) to monitor data quality during acquisition.

Serum analyses were performed on ACQUITY UPLC instruments (Waters Corp., Milford, MA, USA) coupled to Xevo G2-S Q-TOF mass spectrometers (Waters Corp., Manchester, UK) via a Z-spray electrospray ionisation (ESI) source operating in both positive and negative ion modes. Serum datasets consisted of lipid positive (lipid RPC+) and negative (lipid RPC-) and HILIC positive (HILIC+). UPLC-MS acquisitions were structured according to the protocols laid out in Lewis et al. Samples were acquired in batches of up to 1000 study-samples, interleaved with acquisition of alternating pooled QC and LTR samples every five injections (16 per 80 samples), and flanked by a set of dilution series samples to assess linearity of response.

Waters format instrument raw files were converted to .mzNLD for retention-time alignment and feature detection in Progenesis QI (Waters Corp. Milford, MA, USA). Progenesis QI was configured to align retention time to the central LTR sample of the acquisition. Peak detection is configured with a minimum chromatographic peak width of 0.01 minutes, and automatic noise detection set to the minimum threshold of 1. Peaks arising from isotopes and chemical adducts are automatically resolved according to the observed m/z and chromatographic peak-shape, and peaks areas integrated.

Further pre-processing and filtering of UPLC-MS profiling datasets was conducted using the nPYc-Toolbox.[14] Analytical run-order effects were accounted for with an adaption of previously described methods.[35] A robust LOESS regression was generated per-feature, based on the pooled QC samples, in run-order, with the window scaled to include 21 samples. The smoothed response values for each feature were then interpolated to the intermediate study sample injections using simple linear interpolation. Finally, the median intensity of each feature in each analytical batch was aligned.

Extracted features spuriously arising from analytical noise were removed from the dataset by a set of approaches, applied on a per-feature basis. First, dilution series samples were used to assess the linearity of responses of each feature. Detected features were correlated to their expected intensity in the dilution series, and those features showing a Pearson's *r* of less than 0.7 were excluded from further analysis. Second, the relative standard deviation (RSD) of each feature across the study reference samples was calculated, and those features where the RSD exceeded 30% were exclude. Finally, the RSD of each feature across the study samples was calculated (as a proxy for observed biological variance), and those features where this was less than 1.5 times the RSD in the pooled QC samples were excluded.

### Supplementary Figures

**Supplementary Figure 1.** Association of MRI markers and cognition parameters at baseline per 1-SD higher metabolite levels by cohort

#### (a) Lipids

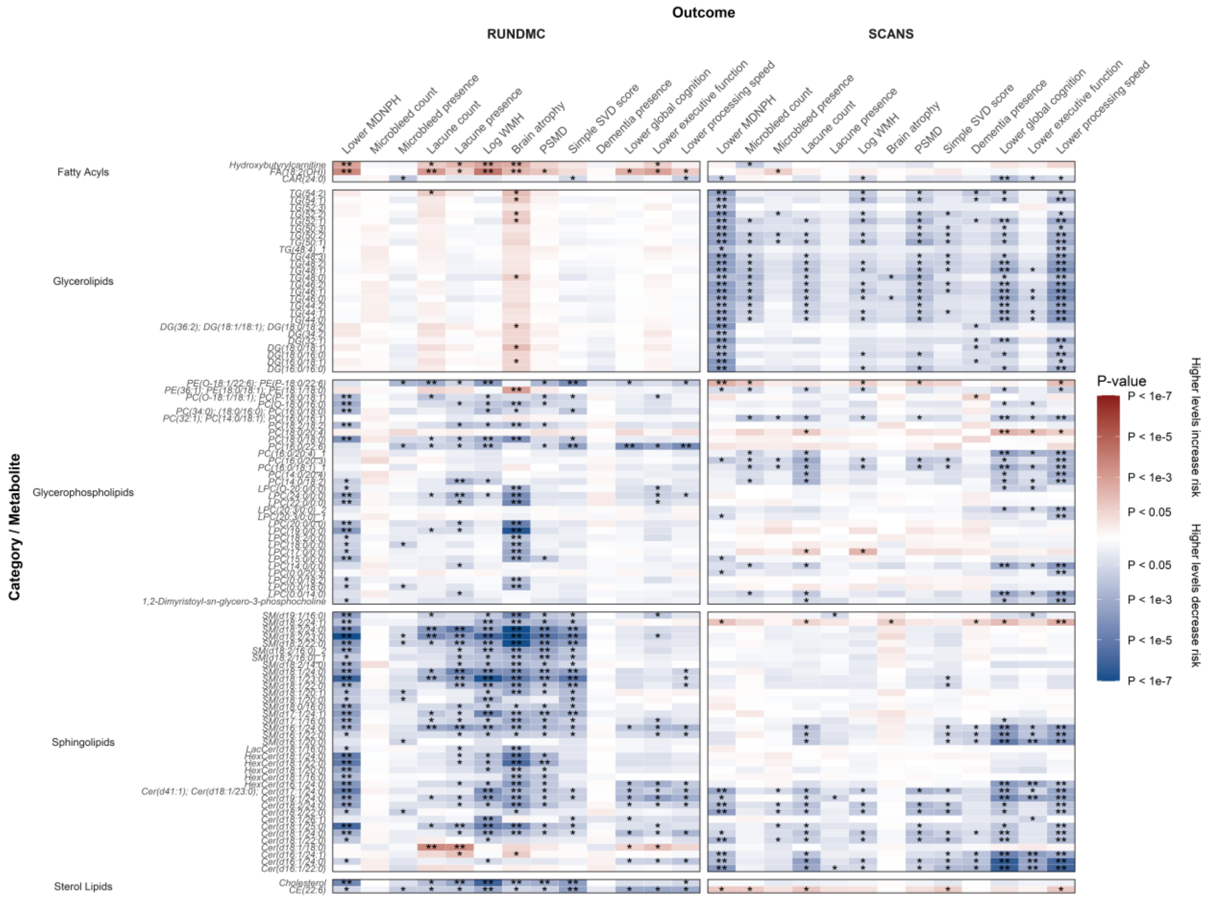

#### (b) Lipoproteins and small molecules

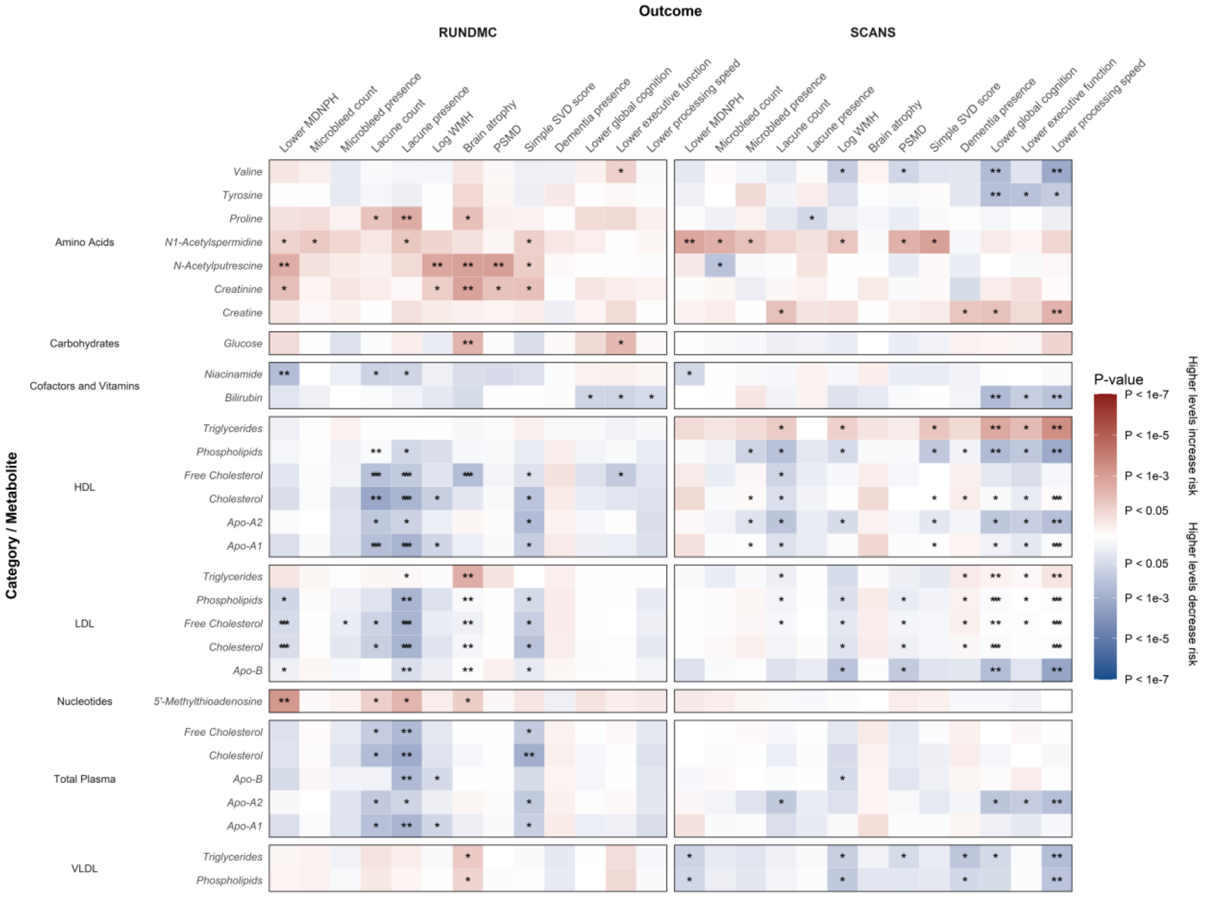


Beta estimates and *P*-values were obtained from linear or logistic regression models adjusted for baseline age and sex and calculated separately within each cohort (RUN-DMC, left; SCANS, right). Colours show magnitude and direction of *P*-value for association of metabolite with each outcome (red indicates positive association and blue indicates inverse association). Analyses were restricted to metabolite-outcome associations that were significant in the primary analysis (**Figure 1**). Asterisks indicate significance: *$P < 0.05$; **FDR $q < 0.05$.
